# Neuropathology-based *APOE* genetic risk score better quantifies Alzheimer’s risk

**DOI:** 10.1101/2022.10.12.22280874

**Authors:** Yuetiva Deming, Eva Vasiljevic, Autumn Morrow, Jiacheng Miao, Carol Van Hulle, Erin Jonaitis, Yue Ma, Vanessa Whitenack, Gwendlyn Kollmorgen, Norbert Wild, Ivonne Suridjan, Leslie M. Shaw, Sanjay Asthana, Cynthia M. Carlsson, Sterling C. Johnson, Henrik Zetterberg, Kaj Blennow, Barbara B. Bendlin, Qiongshi Lu, Corinne D. Engelman, the Alzheimer’s Disease Neuroimaging Initiative

## Abstract

**INTRODUCTION:** *APOE ε4*-carrier status or *ε4* allele count are included in analyses to account for the *APOE* genetic effect on Alzheimer’s disease (AD); however, this does not account for protective effects of *APOE ε2* or heterogeneous effect of *ε2, ε3, ε4* haplotypes.

**METHODS:** We leveraged results from an autopsy-confirmed AD study to generate a weighted risk score for *APOE* (*APOE*-npscore). We regressed cerebrospinal fluid (CSF) amyloid and tau biomarkers on *APOE* variables from the Wisconsin Registry for Alzheimer’s Prevention, Wisconsin Alzheimer’s Disease Research Center, and Alzheimer’s Disease Neuroimaging Initiative (ADNI).

**RESULTS:** The *APOE*-npscore explained more variance and provided a better model fit for all three CSF measures than *APOE ε4*-carrier status and *ε4* allele count. These findings were replicated in ADNI and observed in subsets of cognitively unimpaired participants.

**DISCUSSION:** The *APOE*-npscore reflects the genetic effect on neuropathology and provides an improved method to account for *APOE* in AD-related analyses.

## 1. Introduction

The apolipoprotein E gene (*APOE*) is the predominant genetic risk factor for late-onset Alzheimer’s disease (AD), with three alleles contributing to disease risk: ε2 (reduced risk), ε3 (reference), and ε4 (increased risk). *APOE* genotype is associated with many AD endophenotypes, such as biomarkers reflecting the underlying neuropathology of amyloid plaques and neurofibrillary tangles, such as cerebrospinal fluid (CSF) [1] and positron emission tomography (PET) measures of amyloid and tau [2]. The importance of accounting for the strong genetic effect of *APOE* on AD risk has been recognized in many analyses of AD-related outcomes and researchers often use *APOE ε4*-carrier status (APOE4-status: ε4+/ε4-) [2,3] or, less frequently, the number of *APOE ε4* alleles (*ε4*-count: 0, 1, 2) [4]. Using these methods to model *APOE* genetic risk has limitations: 1. APOE4-status and the *ε4*-count do not account for the effects of reduced risk conferred by *APOE ε2*; 2. an assumption of the allele count approach is that genetic risk of *APOE ε4* is strictly additive; and 3. as a dichotomous variable APOE4-status has limitations in statistical modeling such as loss of statistical power or problems with model convergence [5]. To overcome these limitations, we previously used a weighted score for *APOE* genotype based on risk for AD diagnosis [1,6]. Another group used a similar method to model the *APOE* genetic effect in polygenic risk scores, weighting the number of *APOE ε2* alleles and the number of *APOE ε4* alleles by the effect sizes reported in the Kunkle *et al*., 2019 genome-wide association study [7] for single nucleotide polymorphisms rs7412 (encoding *ε2*) and rs429358 (encoding *ε4*) [8]. One limitation of these *APOE* risk scores is that they are based on clinical diagnosis of AD dementia which can include preclinical AD appearing as controls and dementia cases due to non-AD causes [9–11]. Here we propose an improved method to account for *APOE* genetic risk for AD in statistical analyses using a weighted score based on AD neuropathology, providing a pseudo-continuous variable that does not collapse important genotype categories. By comparing AD cases and controls that were confirmed at autopsy, Reiman *et al*. showed that the odds ratio (OR) has been overestimated for *APOE* ε2/ε2 individuals and underestimated for *APOE* ε4/ε4 individuals in clinical risk studies [12]. Similar results have been obtained using CSF endophenotypes as surrogate measures of AD pathology [13]. We propose that using this *APOE* neuropathology-based score (*APOE*-npscore) will help researchers more accurately account for the genetic effect of *APOE* on the underlying neuropathology of AD. This can increase statistical power, avoid modeling issues that result from low-frequency genotypes with low counts, and allows for a more nuanced variable that may help distinguish AD from diseases with similar clinical appearance.

CSF biomarkers for amyloid-beta1-42 (Aβ42) and amyloid-beta1-42/1-40 (Aβ42/40) ratio and phosphorylated tau 181 (ptau181) are among the gold standard biomarkers for AD. CSF Aβ42 and the Aβ42/40 ratio decrease early in AD and are negatively correlated with amyloid PET [14] and amyloid plaques presence, a hallmark neuropathology of AD [15]. CSF Aβ42/40 ratio has been reported to predict PET amyloid-positivity more accurately than CSF Aβ42 alone, regardless of clinical diagnosis [16,17]. Another key pathological hallmark of AD, neurofibrillary tangles composed of hyperphosphorylated tau, is positively correlated with CSF ptau181 concentration [18]. The ratio of CSF ptau181/Aβ42 is predictive of cognitive decline and conversion to AD dementia [19–21]. These CSF biomarkers change before cognitive symptoms appear [19,22] and can help distinguish AD from other diseases that are clinically similar [23–25]. The correlation with AD neuropathology and measurable changes early in disease made these CSF biomarkers ideal for testing our hypothesis that the *APOE*-npscore is an improvement over other methods in accounting for the *APOE* genetic risk for AD in statistical analyses.

## 2. Methods

### 2.1. Participants

The Institutional Review Boards of all participating institutions approved the study, and research was carried out in accordance with approved protocols. Written informed consent was obtained from participants or their family members. Data were obtained from longitudinal studies of preclinical and clinical AD from the Wisconsin Registry for Alzheimer’s Prevention (WRAP) [26] and the Wisconsin Alzheimer’s Disease Research Center (WADRC). WRAP is a longitudinal observational cohort study, established in 2001, of middle-aged participants that is enriched with people who have a parental history of probable-AD dementia. The WADRC was established in 2009 and is one of the National Institute on Aging (NIA)-designated ADRCs across the United States. Participants enrolled in these studies provided CSF within one year of cognitive testing. Diagnoses were determined by consensus conference of dementia specialists based on NIA-Alzheimer’s Association (NIA-AA) criteria without reference to biomarker status [27,28]. The combined data in these analyses include participants with mild cognitive impairment (MCI), dementia due to suspected AD (dementia-AD), or cognitively unimpaired (CU) individuals.

Data used for replication analyses were obtained from the Alzheimer’s Disease Neuroimaging Initiative (ADNI) database (adni.loni.usc.edu) [29]. The ADNI was launched in 2003 as a public-private partnership, led by Principal Investigator Michael W. Weiner, MD. The primary goal of ADNI has been to test whether serial magnetic resonance imaging (MRI), PET, other biological markers, and clinical and neuropsychological assessment can be combined to measure the progression of MCI and early AD. For up-to-date information, see www.adni-info.org.

### 2.2. Genotyping and scoring

DNA extracted from whole blood samples from WADRC and WRAP participants was genotyped for *APOE ε*2and *APOE ε*4 using competitive allele-specific PCR-based KASP genotyping for rs7412 and rs429358, respectively [6]. Data downloaded from the ADNI database were obtained from DNA extracted from blood, as described previously [30].

To generate the *APOE*-npscore we used a natural log (ln) transformation of the OR values reported in a study of *APOE* genetic risk in autopsy-confirmed AD cases (n = 4018) and controls (n = 989), none of whom were participants in our analyses [12]. OR values were obtained from the Reiman *et al*., 2020, supplementary table which had OR calculated for each *APOE* genotype, using *ε3ε3* as the reference, after adjusting for age and sex: *ε2ε2* OR = 0.16, *ε2ε3* OR = 0.40, *ε3ε3* OR = 1, *ε2ε4* OR = 2.47, *ε3ε4* OR = 5.71, and *ε4ε4* OR = 26.93 [12]. By using the ln(OR), the *APOE*-npscore is negative for haplotypes associated with reduced risk for AD compared to *ε3ε3*, resulting in the *APOE*-npscore: *ε2ε2* = -1.833, *ε2ε3* = -0.916, *ε3ε3* = 0, *ε2ε4* = 0.904, *ε3ε4* = 1.742, and *ε4ε4* = 3.293.

### 2.3. CSF collection and assays

CSF samples from WADRC and WRAP were acquired as described previously [31]. Briefly, samples were collected by lumbar puncture (LP) in the morning after an 8-to 12-hour fast, centrifuged to remove red blood cells or other debris, then 0.5 mL CSF was aliquoted into 1.5-mL polypropylene tubes and stored at -80^∘^C within 30 minutes of collection. WRAP and WADRC CSF samples were assayed at the Clinical Neurochemistry Laboratory, University of Gothenburg under strict quality control procedures. Aβ42, ptau181, and Aβ40 levels in CSF were measured using the Elecsys® β-Amyloid(1-42) CSF, Elecsys Phospho-Tau (181P) CSF, and Elecsys β-Amyloid(1-40) electrochemiluminescence immunoassays, respectively, on the cobas e 601 analyzer (all Roche Diagnostics International Ltd).

ADNI CSF samples, obtained as described in the ADNI procedures manual (http://www.adni-info.org), were also measured using Elecsys CSF immunoassays on a cobas e 601 analyzer at the University of Pennsylvania [32]. ADNI CSF data (versions 2021-01-04 and 2019-07-29) were downloaded in early 2022 from the ADNI database (https://ida.loni.usc.edu) with corresponding participant demographics such as age, sex, diagnosis, and *APOE* genotypes. Data were verified to be the most currently available as of September 7, 2022.

The Elecsys CSF immunoassays have measuring ranges of 200–1700 pg/mL for Aβ42, 0.006– 40.3 ng/mL for Aβ40 (specific for lot used), and 8–120 pg/mL for ptau181 [31,32]. Performance of the assays above these technical limits has not been formally established. Therefore, we only analyzed values within the technical limits. CSF Aβ42/40 and ptau181/Aβ42 ratios were derived from the CSF Aβ42, Aβ40, and ptau181 values.

### 2.4. Statistical analyses

There were 1045 individuals available for analyses in WADRC (n = 380), WRAP (n = 238), and ADNI (n = 427). Initial analyses used WADRC and WRAP combined data, then replication analyses were performed using ADNI data. Statistical analyses were performed in R (version 4.2.0) [33]. Sample characteristics were compared between studies using analysis of variance for continuous measures and chi-square for categorical measures.

Associations between CSF biomarker (Aβ42/40 ratio, ptau181, or ptau181/Aβ42 ratio) and *APOE* variable (*APOE*-npscore, APOE4-status, or ε4-count) were each tested using linear mixed-effects regression in the lmerTest R package (version 3.1-3) [34] with random intercepts for each participant to account for multiple LP visits. Self-reported sex, clinical diagnosis (CU, MCI-AD, dementia-AD), and linear and quadratic terms for mean centered age at LP were entered as fixed covariates. CSF ptau181 values and the ptau181/Aβ42 ratio were ln-transformed and standardized within study. Residual diagnostics, used to verify covariate selection and check model assumptions, were performed using the DHARMa R package (version 0.4.5) [35]. To determine goodness-of-fit and quantify differences in model fit between *APOE*-npscore, APOE4-status (0, 1), and *ε4*-count (0, 1, 2), we compared the Akaike information criterion (AIC), Bayesian Information Criterion (BIC), and the proportion of variance explained by models differing only by the *APOE* variable as predictor. Pseudo-R^2^ statistics were calculated using the MuMIn R package (version 1.47.1) [36] and the marginal R^2^ were compared to determine the difference in variance attributable to the fixed effects portion of each model.

Relative improvement between models was calculated using the ratio of marginal R^2^ values. Other R packages used included kableExtra (version 1.3.4) [37], tableone (version 0.13.2) [38], and sjPlot (version 2.8.11) [39].

## 3. Results

### 3.1. Participant characteristics by study

The characteristics of participants in WADRC, WRAP, and ADNI are shown by study in Table 1. Comparisons between WADRC and WRAP are provided in Supplementary Table S1 and characteristics by ADNI protocol (ADNI1, ADNI2, ADNIGO, and ADNI3) are shown in Supplementary Table S2. Participant characteristics are based on the most recent LP visit; information about multiple LP visits and biomarker values are provided based on longitudinal data (1566 CSF samples). Supplementary Table S3 shows characteristics by study (WADRC, WRAP, and ADNI) for a subset of CU participant samples used in sensitivity analyses.

**Table 1.**
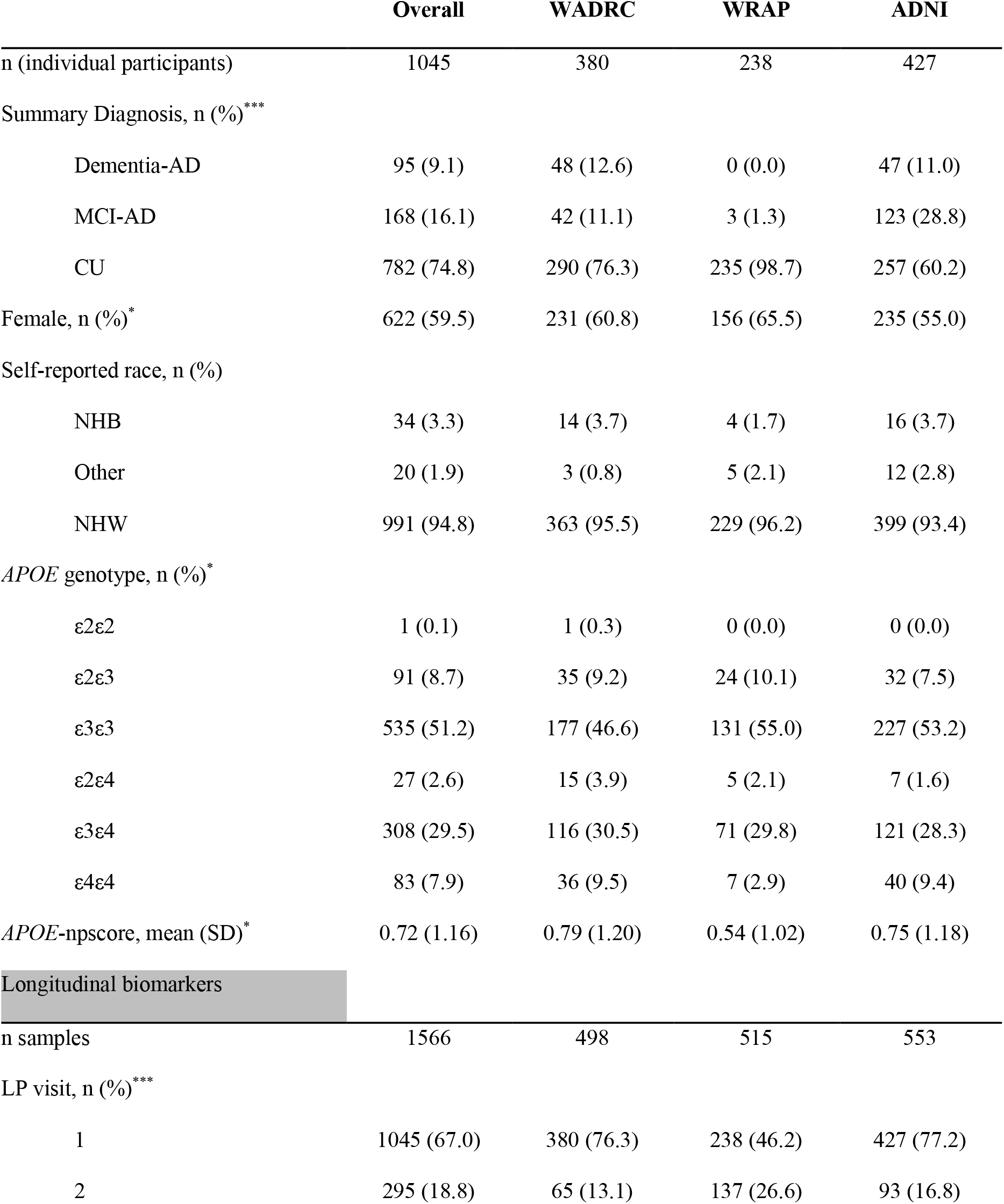
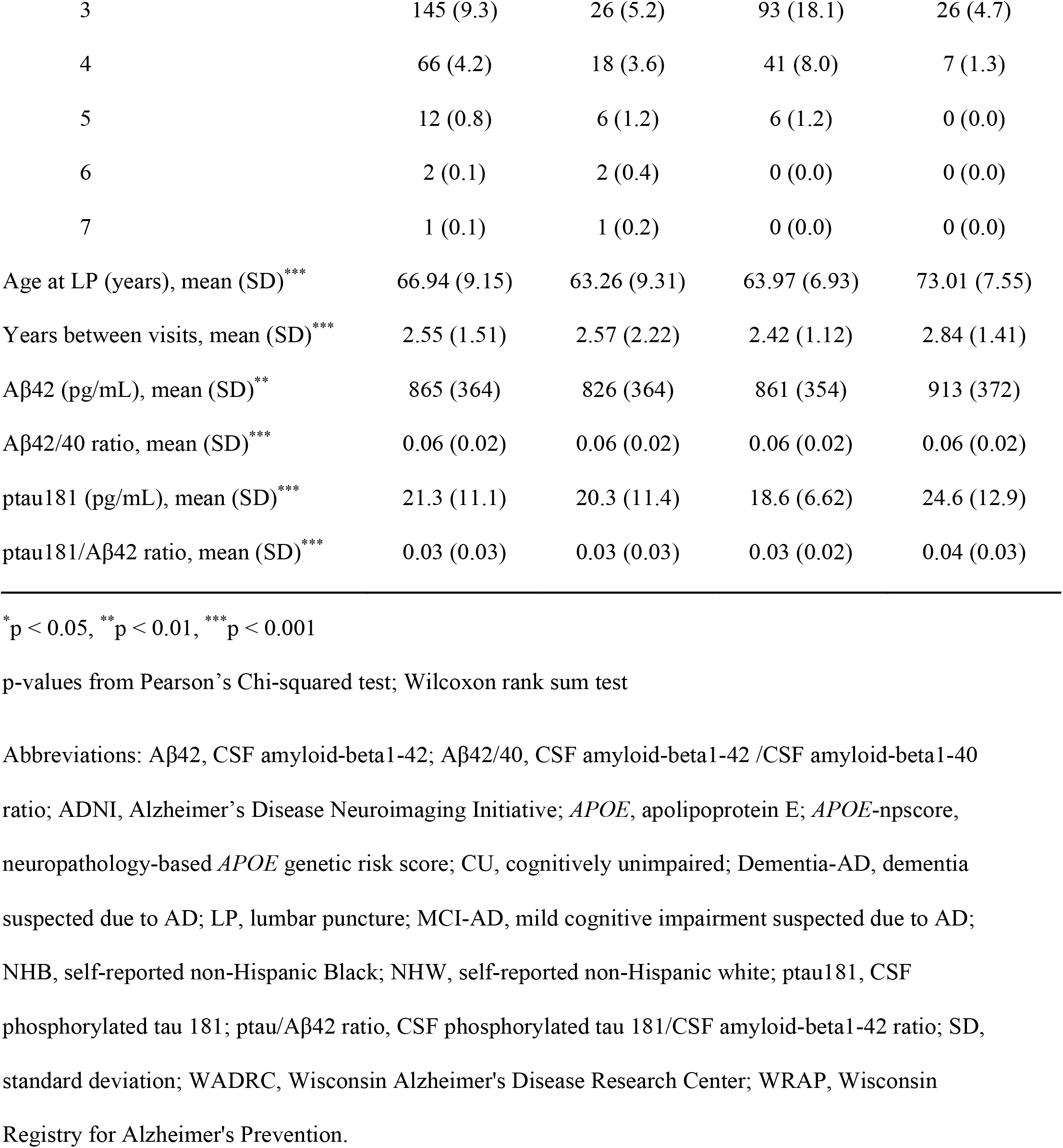
Cohort demographics at most recent lumbar puncture and longitudinal biomarkers

|  | Overall | WADRC | WRAP | ADNI |
| --- | --- | --- | --- | --- |
| n (individual participants) | 1045 | 380 | 238 | 427 |
| Summary Diagnosis, n (%)*** |  |  |  |  |
| Dementia-AD | 95 (9.1) | 48 (12.6) | 0 (0.0) | 47 (11.0) |
| MCI-AD | 168 (16.1) | 42 (11.1) | 3 (1.3) | 123 (28.8) |
| CU | 782 (74.8) | 290 (76.3) | 235 (98.7) | 257 (60.2) |
| Female, n (%)* | 622 (59.5) | 231 (60.8) | 156 (65.5) | 235 (55.0) |
| Self-reported race, n (%) |  |  |  |  |
| NHB | 34 (3.3) | 14 (3.7) | 4 (1.7) | 16 (3.7) |
| Other | 20 (1.9) | 3 (0.8) | 5 (2.1) | 12 (2.8) |
| NHW | 991 (94.8) | 363 (95.5) | 229 (96.2) | 399 (93.4) |
| *APOE* genotype, n (%)* |  |  |  |  |
| ε2ε2 | 1 (0.1) | 1 (0.3) | 0 (0.0) | 0 (0.0) |
| ε2ε3 | 91 (8.7) | 35 (9.2) | 24 (10.1) | 32 (7.5) |
| ε3ε3 | 535 (51.2) | 177 (46.6) | 131 (55.0) | 227 (53.2) |
| ε2ε4 | 27 (2.6) | 15 (3.9) | 5 (2.1) | 7 (1.6) |
| ε3ε4 | 308 (29.5) | 116 (30.5) | 71 (29.8) | 121 (28.3) |
| ε4ε4 | 83 (7.9) | 36 (9.5) | 7 (2.9) | 40 (9.4) |
| *APOE*-npscore, mean (SD)* | 0.72 (1.16) | 0.79 (1.20) | 0.54 (1.02) | 0.75 (1.18) |
| Longitudinal biomarkers |  |  |  |  |
| n samples | 1566 | 498 | 515 | 553 |
| LP visit, n (%)*** |  |  |  |  |
| 1 | 1045 (67.0) | 380 (76.3) | 238 (46.2) | 427 (77.2) |
| 2 | 295 (18.8) | 65 (13.1) | 137 (26.6) | 93 (16.8) |
| 3 | 145 (9.3) | 26 (5.2) | 93 (18.1) | 26 (4.7) |
| 4 | 66 (4.2) | 18 (3.6) | 41 (8.0) | 7 (1.3) |
| 5 | 12 (0.8) | 6 (1.2) | 6 (1.2) | 0 (0.0) |
| 6 | 2 (0.1) | 2 (0.4) | 0 (0.0) | 0 (0.0) |
| 7 | 1 (0.1) | 1 (0.2) | 0 (0.0) | 0 (0.0) |
| Age at LP (years), mean (SD)<sup>***</sup> | 66.94 (9.15) | 63.26 (9.31) | 63.97 (6.93) | 73.01 (7.55) |
| Years between visits, mean (SD)<sup>***</sup> | 2.55 (1.51) | 2.57 (2.22) | 2.42 (1.12) | 2.84 (1.41) |
| Aβ42 (pg/mL), mean (SD)<sup>**</sup> | 865 (364) | 826 (364) | 861 (354) | 913 (372) |
| Aβ42/40 ratio, mean (SD)<sup>***</sup> | 0.06 (0.02) | 0.06 (0.02) | 0.06 (0.02) | 0.06 (0.02) |
| ptau181 (pg/mL), mean (SD)<sup>***</sup> | 21.3 (11.1) | 20.3 (11.4) | 18.6 (6.62) | 24.6 (12.9) |
| ptau181/Aβ42 ratio, mean (SD)<sup>***</sup> | 0.03 (0.03) | 0.03 (0.03) | 0.03 (0.02) | 0.04 (0.03) |

\* $p < 0.05$ , \*\* $p < 0.01$ , \*\*\* $p < 0.001$
p-values from Pearson's Chi-squared test; Wilcoxon rank sum test
Abbreviations: Aβ42, CSF amyloid-beta1-42; Aβ42/40, CSF amyloid-beta1-42 /CSF amyloid-beta1-40 ratio; ADNI, Alzheimer's Disease Neuroimaging Initiative; *APOE*, apolipoprotein E; *APOE*-npscore, neuropathology-based *APOE* genetic risk score; CU, cognitively unimpaired; Dementia-AD, dementia suspected due to AD; LP, lumbar puncture; MCI-AD, mild cognitive impairment suspected due to AD; NHB, self-reported non-Hispanic Black; NHW, self-reported non-Hispanic white; ptau181, CSF phosphorylated tau 181; ptau/Aβ42 ratio, CSF phosphorylated tau 181/CSF amyloid-beta1-42 ratio; SD, standard deviation; WADRC, Wisconsin Alzheimer's Disease Research Center; WRAP, Wisconsin Registry for Alzheimer's Prevention.

All studies were predominantly non-Hispanic white (NHW) and mostly female. There were significant differences in clinical diagnosis across studies. WRAP was comprised of CU individuals with a few MCI-AD, both WADRC and ADNI had similar numbers of dementia-AD, and ADNI had the largest proportion of MCI-AD. Mean *APOE*-npscores were lower in WRAP (0.54 ± 1.02) than WADRC (0.79 ± 1.20) and ADNI (0.75 ± 1.18). Less than 14% of WADRC, almost 27% of WRAP, and less than 17% of ADNI participants had 2 or more LPs with a mean difference ∼2.5 years between visits. Mean age at LP across longitudinal samples was similar in WADRC and WRAP (∼63 years) but older in ADNI (73 years). ADNI participants had significantly higher levels of both CSF Aβ42 (913 ± 372 pg/mL) and ptau181 (24.6 ± 12.9 pg/mL) than WADRC (Aβ42: 826 ± 364 pg/mL; ptau181: 20.3 ± 11.4 pg/mL) and WRAP (Aβ42: 861 ± 354 pg/mL; ptau181: 18.6 ± 6.62 pg/mL). Scatterplots of age at LP against CSF Aβ42, Aβ42/40 ratio, and ptau181 values by study are shown in Figure 1.

**Figure 1.**
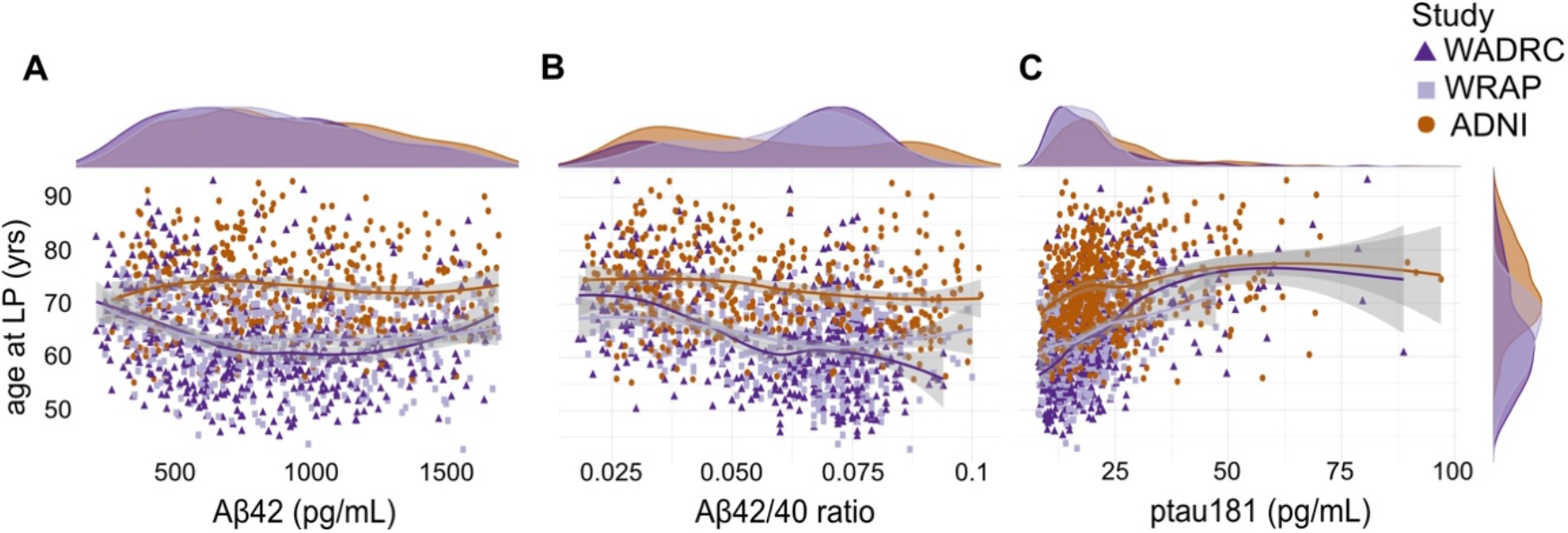
CSF biomarker levels by age at time of lumbar puncture. Scatterplots of the age at time of lumbar puncture (LP) against CSF Aβ42 (A), Aβ42/40 ratio (B), and ptau181 (C) with density plots for each biomarker on top and a density plot for age at LP for all three panels on the right-hand side of (C). Each point represents an individual CSF sample. Densities, loess curves, and points are all color-coded by study as shown in the legend; triangle-shaped points indicate CSF sample data from WADRC, square-shaped points indicate CSF sample data from WRAP, and circle-shaped points indicate CSF sample data from ADNI.

### 3.2. Using ε4-count provided a better model fit than APOE4-status and explained more variance in CSF AD endophenotypes

Some researchers use *ε4*-count instead of the binary APOE4-status, so we tested if *ε4*-count provides a better model fit than APOE4-status. In our preliminary analyses of WADRC and WRAP, APOE4-status and *ε4*-count were associated with CSF Aβ42/40 ratio (β = -0.012, p = 3.98×10^−21^ and β = -0.010, p = 1.04×10^−23^, respectively), ptau181 (β = 0.258, p = 6.49×10^−4^ and β = 0.238, p = 9.20×10^−5^, respectively), and ptau181/Aβ42 ratio (β = 0.524, p = 2.52×10^−14^ and β = 0.460, p = 7.42×10^−17^, respectively); however, models using *ε4*-count explained more variance than APOE4-status with 3.2% relative increase in CSF Aβ42/40 ratio (marginal R^2^ = 0.355 vs 0.344), 2.5% relative increase in ptau181 (marginal R^2^ = 0.208 vs 0.203), and 3.4% increase in ptau181/Aβ42 ratio (marginal R^2^ = 0.370 vs 0.358). Model fit comparisons are shown in Table 2 and detailed regression results in Supplementary Tables S4-S6.

**Table 2.** Linear-mixed effects model comparisons between *APOE* genetic variables.

| | CSF Aβ42/40 ratio | | | | CSF ptau181 | | | | CSF ptau181/Aβ42 ratio | | | |
| --- | --- | --- | --- | --- | --- | --- | --- | --- | --- | --- | --- | --- |
|  | n | R<sup>2</sup> | AIC | BIC | n | R<sup>2</sup> | AIC | BIC | n | R<sup>2</sup> | AIC | BIC |
| WADRC/WRAP |  |  |  |  |  |  |  |  |  |  |  |  |
| APOE4-status | 948 | 0.344 | -5728 | -5684 | 971 | 0.203 | 1641 | 1685 | 924 | 0.358 | 1781 | 1825 |
| ε4-count | 948 | 0.355 | -5740 | -5696 | 971 | 0.208 | 1637 | 1681 | 924 | 0.370 | 1770 | 1813 |
| *APOE*-npscore | 948 | **0.360** | **-5746** | **-5702** | 971 | **0.210** | **1635** | **1679** | 924 | **0.378** | **1761** | **1805** |
| ADNI |  |  |  |  |  |  |  |  |  |  |  |  |
| APOE4-status | 423 | 0.315 | -2285 | -2248 | 548 | 0.200 | 1156 | 1195 | 418 | 0.344 | 909 | 945 |
| ε4-count | 423 | 0.359 | -2309 | -2272 | 548 | 0.208 | 1152 | 1190 | 418 | 0.394 | 880 | 917 |
| *APOE*-npscore | 423 | **0.375** | **-2318** | **-2282** | 548 | **0.218** | **1147** | **1186** | 418 | **0.409** | **872** | **908** |

**optimal model fit criteria (largest marginal R<sup>2</sup>, smallest AIC, and smallest BIC) bolded**
Abbreviations: Aβ42/40, CSF amyloid-beta1-42/amyloid-beta1-40 ratio; ADNI, Alzheimer's Disease Neuroimaging Initiative; AIC, Akaike Information Criterion; *APOE*-npscore, *APOE* neuropathology-based score; APOE4-status, *APOE4*-carrier status (0, 1); BIC, Bayesian Information Criterion; ε4-count, number of *APOE* ε4 alleles (0, 1, 2); CSF, cerebrospinal fluid; n, number of observations; ptau181, CSF phosphorylated tau 181; ptau/Aβ42 ratio, CSF phosphorylated tau 181/CSF amyloid-beta1-42 ratio; R<sup>2</sup>, marginal R<sup>2</sup> (variation explained by fixed effects); WADRC, Wisconsin Alzheimer's Disease Research Center; WRAP, Wisconsin Registry for Alzheimer's Prevention.

As shown in Table 2, our findings were replicated in ADNI. Models with *ε4*-count explained more variance than APOE4-status with 14% relative increase explained in CSF Aβ42/40 ratio (marginal R^2^ = 0.359 vs 0.315), 4% increase in ptau181 (marginal R^2^ = 0.208 vs 0.200), and 14.5% increase in ptau181/Aβ42 ratio (marginal R^2^ = 0.394 vs 0.344). Detailed results are shown in Supplementary Tables S7-S9.

### 3.3. APOE-npscore provided a better model fit than ε4-count and explained more variance in CSF AD endophenotypes

After determining that ε4-count provided a better model fit than APOE4-status, we tested if the *APOE*-npscore improved the model fit even more than *ε4*-count. In the WADRC and WRAP, *APOE*-npscore was associated with CSF Aβ42/40 ratio (β = -0.006, p = 6.63×10^−25^), ptau181 (β = 0.140, p = 2.35×10^−5^), and ptau181/Aβ42 ratio (β = 0.264, p = 1.09×10^−18^). Models using *APOE*-npscore explained more variance than *ε4*-count with a relative increase of 1.4% variance explained in CSF Aβ42/40 ratio (marginal R^2^ = 0.360 vs 0.355), 1% increase in ptau181 (marginal R^2^ = 0.210 vs 0.208), and 2.2% increase explained in ptau181/Aβ42 ratio (marginal R^2^ = 0.378 vs 0.370). As shown in Table 2, both AIC and BIC model selection criteria consistently supported models using the *APOE*-npscore over models using the *ε4*-count and models using the APOE4-status. Detailed results are shown in Supplementary Tables S4-S6.

Our findings were replicated in the ADNI. *APOE*-npscore explained more variance than *ε4*-count with a relative increase of 4.5% explained in CSF Aβ42/40 ratio (marginal R^2^ = 0.375 vs 0.359), 4.8% increase in ptau181 (marginal R^2^ = 0.218 vs 0.208), and 3.8% increase in ptau181/Aβ42 ratio (marginal R^2^ = 0.409 vs 0.394). Detailed results are shown in Supplementary Tables S7-S9.

### 3.4. APOE-npscore provided a better model fit in subset of cognitively unimpaired participants

Previous studies have reported preclinical effects of APOE4-status and *ε4*-count on CSF AD biomarkers [40,41]. To test if the *APOE*-npscore could provide a better model fit than APOE4-status and *ε4*-count when analyzing CSF AD endophenotypes before cognitive symptoms appear, we reran the analyses in a subset of longitudinal samples from participants who were CU at LP (described in Supplementary Table S3). There were significant associations between CSF Aβ42/40 ratio and the *APOE*-npscore (β = -0.005, p = 1.06×10^−18^), APOE4-status (β = -0.011, p = 1.47×10^−16^), and *ε4*-count (β = -0.010, p = 4.63×10^−18^); between ptau181 and *APOE*-npscore (β = 0.110, p = 1.52×10^−3^), APOE4-status (β = 0.217, p = 4.68×10^−3^), and *ε4*-count (β = 0.191, p = 3.32×10^−3^); and between ptau181/Aβ42 ratio and *APOE*-npscore (β = 0.241, p = 1.17×10^−13^), APOE4-status (β = 0.491, p = 1.04×10^−11^), and *ε4*-count (β = 0.437, p = 7.85×10^−13^). There was also more variance explained by the *APOE*-npscore than the APOE4-status with a relative increase of 3.7% in CSF Aβ42/40 ratio (marginal R^2^ = 0.222 vs 0.214), 1.4% in CSF ptau181 (marginal R^2^ = 0.150 vs 0.148), and 4.9% in CSF ptau181/Aβ42 ratio (marginal R^2^ = 0.216 vs 0.206). *APOE*-npscore also explained more variance than *ε4*-count with a relative increase of 1% in CSF Aβ42/40 ratio (marginal R^2^ = 0.222 vs 0.220) and 1.4% in ptau181/Aβ42 ratio (marginal R^2^ = 0.216 vs 0.213), but no difference in ptau181 (marginal R^2^ = 0.150 vs 0.150). Both AIC and BIC model selection criteria consistently supported models using the *APOE*-npscore (Aβ42/40 ratio: AIC = -5175, BIC = -5137; ptau181: AIC = 1304, BIC = 1342; ptau181/Aβ42 ratio: AIC = 1512, BIC = 1550) over models using the *ε4*-count (Aβ42/40 ratio: AIC = -5172, BIC = -5134; ptau181: AIC = 1305, BIC = 1343; ptau181/Aβ42 ratio: AIC = 1516, BIC = 1553) and models using the APOE4-status (Aβ42/40 ratio: AIC = -5165, BIC = -5127; ptau181: AIC = 1306, BIC = 1344; ptau181/Aβ42 ratio: AIC = 1521, BIC = 1558). Detailed results are shown in Supplementary Tables S10-S12.

These findings were replicated in a subset of samples from participants who were CU in ADNI (described in Supplementary Table S3). CSF Aβ42/40 ratio was associated with the *APOE*-npscore (β = -0.009, p = 8.13×10^−12^), APOE4-status (β = -0.018, p = 5.94×10^−9^), and *ε4*-count (β = -0.016, p = 6.78×10^−11^). CSF ptau181 was associated with *APOE*-npscore (β = 0.181, p = 3.74×10^−4^), APOE4-status (β = 0.324, p = 4.04×10^−3^), and *ε4*-count (β = 0.298, p = 1.54×10^−3^).

The CSF ptau181/Aβ42 ratio was associated with *APOE*-npscore (β = 0.335, p = 8.55×10^−11^), APOE4-status (β = 0.602, p = 3.26×10^−7^), and *ε4*-count (β = 0.583, p = 8.93×10^−10^). There was more variance explained by the *APOE*-npscore than the APOE4-status with a relative increase of 20.5% in CSF Aβ42/40 ratio (marginal R^2^ = 0.264 vs 0.219), 9% in CSF ptau181 (marginal R^2^ = 0.182 vs 0.167), and 22.5% in CSF ptau181/Aβ42 ratio (marginal R^2^ = 0.299 vs 0.244). *APOE*-npscore also explained more variance than *ε4*-count with a relative increase of 6.5% in CSF Aβ42/40 ratio (marginal R^2^ = 0.264 vs 0.248), 5.8% in ptau181 (marginal R^2^ = 0.182 vs 0.172), and 6% in ptau181/Aβ42 ratio (marginal R^2^ = 0.299 vs 0.282). Both AIC and BIC model selection criteria consistently supported models using the *APOE*-npscore (Aβ42/40 ratio: AIC = -1422, BIC = -1387; ptau181: AIC = 657, BIC = 696; ptau181/Aβ42 ratio: AIC = 508, BIC = 543) over models using the *ε4*-count (Aβ42/40 ratio: AIC = -1418, BIC = -1382; ptau181: AIC = 660, BIC = 699; ptau181/Aβ42 ratio: AIC = 513, BIC = 548) and models using the APOE4-status (Aβ42/40 ratio: AIC = -1409, BIC = -1373; ptau181: AIC = 662, BIC = 701; ptau181/Aβ42 ratio: AIC = 525, BIC = 560). Detailed results are shown in Supplementary Tables S13-S15.

## 4. Discussion

We report here a method for translating *APOE* haplotype (ε2ε2, ε2ε3, ε2ε4, ε3ε3, ε3ε4, and ε4ε4) into a pseudo-continuous measure reflecting *APOE* genetic risk for autopsy-confirmed AD. We demonstrated the *APOE*-npscore provides a better model fit than dichotomous APOE4-status, and even *ε4*-count, in statistical models of CSF endophenotypes for AD neuropathology (Aβ42/40 ratio, ptau181, and ptau181/Aβ42 ratio). Although some of the statistical improvements we reported in this study were small, there are several benefits of using *APOE*-npscore in AD-related research. Not only does it more accurately represent corresponding risk for each haplotype, the *APOE*-npscore more closely reflects the genetic effect of *APOE* on AD neuropathology. It allows researchers to account for effects of ε2 and ε4 in one variable. As an improvement to *APOE* clinical risk scores, by using the results of a large autopsy-confirmed AD case-control study, [12] we minimize bias from misclassified dementia cases and preclinical controls.

CSF biomarkers change years before cognitive symptoms appear and are not only correlated with AD neuropathology but also with *APOE* [1,22,42]. As expected, all the methods we tested for encoding *APOE* genotype were significantly associated with CSF Aβ42/40 ratio, ptau181, and ptau181/Aβ42 ratio. In all three AD endophenotypes, *APOE*-npscore appeared to provide a better model fit than APOE4-status and *ε4*-count and there was more variance explained by the *APOE*-npscore models, consistently observed in WADRC, WRAP, and ADNI as well as subsets of CU individuals.

There are important limitations in this study to consider for future research. The Reiman *et al*. study that provided OR used to derive the *APOE*-npscore consisted of NHW individuals [12] and the cohorts included in our analyses comprised >94% NHW participants. With reported disparities in biomarker outcomes and in *APOE* genetic risk, there is evidence our findings may not translate directly to other populations [43-54]. Self-identified non-Hispanic Black (NHB) individuals are at a greater risk of AD dementia than NHW; however, even though *APOE* ε4 is more common in African genetic ancestry (rs429358 MAF: 0.26 in AFR vs. 0.14 in EUR), studies suggest *APOE* ε4 has a weaker effect, or no effect, on AD dementia in NHB individuals [45–49]. Linkage disequilibrium structure of the *APOE* gene region varies across genetic ancestries [50] and studies show there are genetic haplotypes predominantly present in African genetic ancestry that may explain some of the differences in *APOE* genetic effect on AD risk [3,51]. Disparities in the *APOE* ε4 genetic effect on AD risk have been observed in other populations. Although studies of Chinese patients show that *APOE* ε4 increases risk for AD similar to NHW [52,53], a study of neuroimaging and cognitive testing from 811 American Indians in the Strong Heart Study found no evidence of increased risk from *APOE* ε4 [54].

Racial disparities in these examples and other AD biomarker studies demonstrate that although race is not a biological construct, the biological outcomes of interest are confounded by racial disparities in study recruitment and research [55,56]. Further research with diverse cohorts will be necessary to test and adapt the *APOE*-npscore to be useful for a much broader group of people, many of whom are at an even greater risk for AD dementia than NHW individuals.

## Data Availability

All Alzheimer's Disease Neuroimaging Initiative data are available through the LONI Imaging & Data Archive. Interested scientists may apply for access on the Alzheimer's Disease Neuroimaging Initiative website (http://adni.loni.usc.edu/data-samples/access-data/). Interested scientists may apply to access data from the Wisconsin Alzheimer's Disease Research Center and Wisconsin Registry for Alzheimer's Prevention through the website (https://www.adrc.wisc.edu/apply-resources).

https://ida.loni.usc.edu

https://www.adrc.wisc.edu/apply-resources

## Abbreviations

Aβ42: amyloid-beta1-42
Aβ42/40: amyloid-beta1-42 to amyloid-beta1-40 ratio
AD: Alzheimer’s disease
ADNI: Alzheimer’s Disease Neuroimaging Initiative
AIC: Akaike information criterion
APOE: Apolipoprotein
E: APOE4-status:
*APOE ε4*: carrier status (1 = if at least one *APOE ε4* allele, 0 = if none)
*APOE*-npscore: weighted *APOE* genetic risk score based on autopsy-confirmed
AD, BIC: Bayesian Information Criterion
CSF: cerebrospinal fluid
CU: cognitively unimpaired
dementia-AD: dementia suspected due to
AD: *ε4*-countnumber of *APOE ε4* alleles (0, 1, 2)
ln: natural log
LP: lumbar puncture
MCI: mild cognitive impairment
MRI: magnetic resonance imaging
NIA: National Institutes on Aging
NHW: non-Hispanic white
NHB: non-Hispanic Black
OR: odds ratio
PET: positron emission tomography
ptau181: phosphorylated tau 181
ptau181/Aβ42: phosphorylated tau 181 to amyloid-beta1-42 ratio
WADRC: Wisconsin Alzheimer’s Disease Research Center
WRAP: Wisconsin Registry for Alzheimer’s Prevention

## Acknowledgements

We would like to thank the WRAP and WADRC research participants and their families for their generosity; none of this research would be possible without them. We also would like to thank the staff of the WRAP and WADRC for all of their hard work. We appreciate the ADNI investigators and the contribution of the late John Q. Trojanowski, as co-leader of the ADNI Biomarker Core at the University of Pennsylvania, for providing data for replication. CSF assay kits were provided by Roche Diagnostics GmbH, Penzberg, Germany.

COBAS, COBAS E, and ELECSYS are trademarks of Roche. The Elecsys β-Amyloid(1−42) CSF and Phospho-Tau (181P) CSF immunoassays are approved for clinical use in CE-mark accepting countries. The robust prototype β-Amyloid(1−40) CSF immunoassay is for investigational use only and not commercially available.

## Data availability

All ADNI data are available through the LONI Imaging & Data Archive. Interested scientists may apply for access on the ADNI website (http://adni.loni.usc.edu/data-samples/access-data/). Interested scientists may apply to access data from the WADRC and WRAP through the website (https://www.adrc.wisc.edu/apply-resources).

